# Profile of Mucormycosis Cases from a Network of Hospitals in North India Amidst COVID-19 Pandemic

**DOI:** 10.1101/2021.08.25.21262404

**Authors:** Sandeep Budhiraja, Mona Aggarwal, Monica Mahajan, Abhaya Indrayan, Vinita Jha, Ambrish Mittal, Sanjay Sachdeva, Sumit Mrig, Anurag Jain, Ravinder Gera, Rahul Aggarwal, Suven Kalra, WVPS Ramalingam, Anupal Deka, Arjun Das, D Jijina, Ajit Mansingh, Anuj Singhal, Rajashekhar Reddi, Puneet Aggarwal, Mukesh Kumar, JD Mukherjee, Vivek Nangia, Ajay Lall, Omender Singh, Arun Dewan, Ajay Jain, Gita G Shrivastava, Mala Bhattacharjee, Meena Nihalani, Manoj Kumar, Meenakshi Jain, Mukesh Mehra, Vijay Arora, Viresh Prashant Mehta, Dilip Bhalla, Amit Batra, Rajesh Gupta, Vivek Kumar, Sanjeev Dua, Praveen Pandey, Y P Singh, Mohit Mathur, Ashok Singh, Sanjeev Arora, Ajay Kumar Gupta, Pankaj Nand Choudhary, Manoj Singh, Namita Kaul, Sitla Prasad Pathak, Sharad Joshi, Manish Gupta, Rajesh Mishra, Alok Joshi, Manoj Aggarwal, Rajiv Gupta, Vandana Boobna, Yogesh Kumar Chhabra, Inder Mohan Chugh, Sandeep Garg, Vikas Mittal, Neha Sood, Anil Kumar, Rajesh Kumar Pande, V P Singh, Iram Khan, Nitin Garg, Puneet Tyagi, Shantanu Belwal, Anup Kumar Roy, Deepak Bhasin, Sachin Pandove, Ravikant Bahl, Prateek Soni

## Abstract

Incidence of mucormycosis suddenly surged in India after the second wave of COVID-19. This is a crippling disease and needs to be studied in detail to understand the disease, its course, and the outcomes.

Between 1^st^ March and 15^th^ July 2021, our network of hospitals in North India received a total of 155 cases of COVID-associated mucormycosis cases as all of them reported affliction by COVID-19 earlier or concurrent. Their records were retrieved from the Electronic Health Records system of the hospitals and their demographics, clinical features, treatments, and outcomes were studied. More than 80% (125 cases) had proven disease and the remaining 30 were categorized as ‘possible’ mucormycosis as per the EORTC criteria.

More than two-thirds (69.0%) of the cases were males and the mean age was 53 years for either sex. Nearly two-thirds (64.5%) had symptoms of nose and jaws and 42.6% had eye involvement. Some had multiple symptoms. As many as 78.7% had diabetes and 91.6% gave history of use of steroids during COVID-19 treatment. The primary surgery was functional endoscopic sinus surgery (FESS) (83.9%). Overall mortality was 16.8%, which is one-and-a-half times the mortality in hospitalized COVID-19 patients in the corresponding population. Occurrence of mucormycosis was associated with diabetes and use of steroids, but mortality was not associated with either of them. Cases undergoing surgery and on antifungal had steeply lower mortality (11.9% vs. 50.0%, P < 0.001) than those who were exclusively on antifungal drugs. Treatment by different drugs did not make much of a difference in mortality.

## Introduction

In the pre-COVID era, mucormycosis was labelled as a rare and emerging fungal infection. Inspite of this, India has had the highest burden of mucormycosis globally, with an estimated burden of 140 cases per million population [1]. Suddenly, mucormycosis has become a scourge in post COVID-19 patients and India has seen a deluge of so called ‘black-fungus’ patients after the second wave of COVID-19.

We were exposed to this ubiquitous, saprophytic, filamentous mold on a daily basis. In the moderate and severe cases of COVID-19, the immune system is severely compromised, leading to a severe form on angio-invasive COVID-19 associated mucormycosis (CAM) which has a mortality as high as 80% if a patient goes untreated, or remains untreated long and even after treatment, mortality still could be 40-50% [2]. The fungus thrives in uncontrolled diabetics with ketoacidosis as well as in patients with recent steroid use. An acidic pH increases free iron levels in serum (hyperferritinemia) which promotes fungal growth. Macrophage and neutrophil function are impaired in diabetics and lymphopenia in infection with SARS-CoV2 proves to be the precipitating factor for total breakdown of immune machinery. The triad of hyperglycemia, steroid use and acidosis promotes fungal spores to germinate. The most plausible explanation for this recent surge in mucormycosis cases is believed to be the unparalleled, irrational and prolonged use of steroids in COVID-19 patients. The delta variant of the SARS-CoV-2 virus and prolonged ICU stay maybe another reason for higher cases of mucormycosis. Use of industrial oxygen in the wake of an acute shortage of oxygen in hospitals may have contributed to this huge case load [3].

The spectrum of syndromes associated with mucormycosis include rhinocerebral, pulmonary, gastrointestinal, cutaneous, and disseminated disease. Rarer manifestations include renal mucormycosis and prosthetic valve endocarditis. Rhino-orbital-cerebral mucormycosis is the commonest presentation. Treatment of *Mucorales* involves aggressive surgery, antifungal treatment with Amphotericin B, posaconazole or isavuconazole and correction of the underlying cause. Withdrawal of steroids and immunosuppressants can be life-saving. Avoiding overzealous use of antibiotics, zinc and iron supplements, steam inhalation, and voriconazole prophylaxis are advisable.

An early diagnosis and a multi-disciplinary orchestrated guidelines based management is key to survival. On 10th May, 2021 the Government of India declared CAM as a ‘Mucormycosis epidemic’ in view of sudden surge in cases in the second wave. Until 19 May 2021, approximately 5500 people were affected with CAM in India, resulting in 126 casualties [4]. We report here, the clinical profile of all COVID-19 associated mucormycosis cases admitted in eleven network hospitals spread across 5 states of North India, during Wave-2 (March to July 2021).

## Material and Methods

This is a retrospective, observational, multi-centre study that included all the cases recorded as mucormycosis either at discharge or at death, between 1st March 2021 and 15th July 2021 in our network of hospitals in North India. Their records were retrieved from the Electronic Health Records system and their demographic and clinical profile, the hospital course, and the outcome were noted. All these cases gave history of COVID-19, diagnosed by RT-PCR for SARS-CoV-2 or by an alternative method like rapid antigen test, IgG neutralizing antibody to SARS-CoV-2 or a high-resolution CT scan of chest. Thus, all the cases were of COVID associated mucormycosis (CAM). The day of COVID diagnosis was recorded in the data sheet.

The patients were classified into Proven, Probable or Possible Mucormycosis as per the case definition suggested by the European Organization for Research and Treatment of Cancer (EORTC) and the National Institute of Allergy and Infectious Diseases Mycoses Study Group (MSG) [5]. These definitions are as follows:

### Proven Invasive Mucormycosis

Confirmed by pathology and/or culture.

### Probable Invasive Mucormycosis

Probable disease requires a host factor, a clinical criterion, and a mycological criterion. Host factors include neutropenia, allogeneic stem cell transplant, prolonged use of corticosteroids, and treatment with other recognized T cell immunosuppressants. Clinical criteria include lower respiratory tract disease, tracheobronchitis, sinonasal infection and CNS infection. Mycological criteria include mold in sputum, bronchoalveolar lavage fluid, bronchial brush, or sinus aspirate.

### Possible Invasive Mucormycosis

Possible disease requires only a host factor and a clinical criterion from Probable Invasive Mucormycosis.

The characteristics studied were age, sex, symptoms, disease severity, and presence of diabetes. Details of COVID-19 disease in terms of diagnostic modality, place of treatment (home or hospital), need of oxygen and use of steroids were also recorded. The interval between diagnosis of COVID-19 and admission to hospital for suspected mucormycosis was calculated. The records of their ferritin, CRP levels and radiological imaging (CT, MRI) where ever available, were also extracted. The hospital course for the need of oxygen and mechanical ventilation, and treatment in terms of usage of antifungal (amphotericin B and posaconazole), antibiotics, and surgery was also retrieved. The diagnosis was finally confirmed from the histopathological report of the surgical specimen in the cases who underwent surgery. The outcome under study was in-hospital mortality. Whenever possible, we compared the outcomes of CAM patients with recovered COVID-19 patients without any superadded infection, reported for cases admitted during the second wave from the comparable population data set.

Statistical significance of the difference in mortality was evaluated by chi-square test. Wherever the expected cell frequency was less than 5, Fisher exact test was used. Trend in mortality over increasing age was evaluated by Cochran’s test for trend. For comparison in the case of highly skewed distributions, Mann-Whitney test was used. The significance level was set at 5%. SPSS 21 was used for all the calculations.

### Ethics Committee Approval and Consent

This study was approved by the Institutional Ethics Committee, Max Super Speciality Hospital (A unit of Devki Devi Foundation), Address : Service Floor, Office of Ethics Committee, East Block, next to Conference Room, Max Super Speciality Hospital, Saket (A unit of Devki Devi Foundation), 2, Press Enclave Road, Saket, New Delhi – 110017 vide ref. no. BHR/RS/MSSH/DDF/SKT-2/IEC/IM/21-19 dated 19th July2021. The IEC provided no objection and approved the publication of this manuscript.

All the admitted patients gave a prior consent for their anonymised data to be used for research purpose.

## Results

### Characteristics and Hospital Course of the Cases

A total of 155 cases of mucormycosis were discharged or died in our network during the period of the study. More than two-thirds (69.0%) were males (Table 1). The mean age of the patients was 53.2 years (SD = 12.2). Males had an average age of 53.0 years (SD = 12.8) and females 53.6 years (SD = 10.7). Overall age-distribution in the two sexes was not statistically significantly different (P = 0.435).

**Table 1.** Demographic and clinical characteristics of the cases.

| Characteristics | Number | Percent |
| --- | --- | --- |
| Total Cases | 155 | 100.0 |
| Demographics |  |  |
| Total Cases | 155 | 100.0 |
| Mean age in years (SD) |  | 53.2 (12.2) |
| Male | 107 | 69.0 |
| Mean age in years (SD) |  | 53.0 (12.8) |
| Female | 48 | 31.0 |
| Mean age in years (SD) |  | 53.6 (10.7) |
| Symptoms |  |  |
| Eye | 66 | 42.6 |
| Pain | 22 | 14.2 |
| Swelling (Proptosis) | 34 | 21.9 |
| Blurring of vision | 31 | 20.0 |
| Drooping of eyelids | 2 | 1.3 |
| Nose and jaw | 100 | 64.5 |
| Nasal bleed | 16 | 10.3 |
| Stuffiness & congestion | 33 | 21.3 |
| Jaw pain/ toothache | 14 | 9.00 |
| Facial pain/ paraesthesia | 57 | 36.8 |
| Pulmonary | 27 | 17.4 |
| Cough | 20 | 12.9 |
| SOB Dyspnoea | 16 | 10.3 |
| Others | 40 | 25.8 |
| Headache | 35 | 22.6 |
| Seizures | 1 | 0.6 |
| Weakness | 4 | 2.6 |
| Fever | 33 | 21.3 |
| Vomiting and loose stools | 4 | 2.6 |
| Length of stay in the hospital (days) |  |  |
| Ward | 110 | 71.0 |
| Mean length of stay in ward (SD) |  | 14.1 (10.9) |
| ICU | 45 | 29.0 |
| Mean length of stay in ICU (SD) |  | 16.9 (10.8) |
| Radiological Investigations |  |  |
| CT/MRI | 140 | 90.3 |
| CT & MRI | 16 | 10.3 |
| CT | 49 | 31.6 |
| MRI | 75 | 48.4 |
| Details of Surgery |  |  |
| Surgery | 135 | 87.1 |
| FESS | 130 | 83.9 |
| Orbital decompression | 21 | 13.5 |
| Orbital exenteration | 7 | 4.5 |
| Maxillectomy | 11 | 7.1 |
| Craniotomy | 2 | 1.3 |

Commonest symptom (64.5%) in these cases was related to nose and jaw, most (36.8%) of these had facial pain and paraesthesia (Table 1). The next common symptom was related to eyes, reported by 42.6% cases. Eye symptoms included pain (14.2%), swelling (21.9%), blurring of vision (20.0%), and drooping of eyelids (1.3%). Nearly one-sixth (17.4%) had pulmonary symptoms such as cough and shortness of breath. Among other symptoms, headache was reported by 22.6% and fever by 21.3%.

Forty five (29.0%) cases were admitted in ICU, where they had a mean length of stay 16.9 days (SD = 10.8) and the rest (71%) were treated in wards with mean length of stay was 14.1 days (SD = 10.9) (P=_0.148) (Table 1). Surgery was performed in 135 (87.1%) cases, predominantly functional endoscopic sinus surgery (FESS) (83.9%). Many had multiple surgeries. Orbital decompression (13.5%), exenteration (4.5%), maxillectomy (7.1%) and craniotomy (1.3%) were the other surgical procedures performed in these patients (Table 1). The other 20 patients (12.9%) did not undergo any surgery as decided by the treating surgeon. Ten of these patients were very sick, including 5 on mechanical ventilator and ultimately died. Such cases were deemed unfit for surgery due to extremely high risk to life. The remaining 10 cases not operated had mild illness and the decision was to treat them with only antifungals (Amphotericin B and/or Posaconazole). CT (Paranasal Sinuses) or MRI (paranasal sinuses with orbits and brain) was done in a total of 140 (90.3%) cases. Further details are in Table 1.

### Characteristics of the Patients During Their Acute COVID-19 Illness Phase

During the acute COVID-19 infection, more than one-fourth (26.4%) were treated at their home and the remaining 73.6% at the hospital (Table 2). As many as 91.6% reported that they received steroids during their COVID treatment and 63.8% received oxygen at that time. More than three-fourths (78.7%) had type 2 diabetes mellitus (Table 2). Eighteen (11.6%) had concurrent mucormycosis along with COVID-19 (presented with mucormycosis within 24-48 hours of diagnosis of COVID-19) but the remaining 88.4% developed mucormycosis and got admitted after a median interval of 22 days (IQR 15 – 31), with the longest interval being 69 days from the diagnosis of COVID-19.

**Table 2.** Characteristics of patients when first diagnosed with COVID-19.

| Particulars | n = 155 | Number | Percent |
| --- | --- | --- | --- |
| Place of treatment | Home | 41 | 26.4 |
|  | Hospital | 114 | 73.6 |
| COVID diagnosis | RT-PCR+ | 131 | 84.5 |
|  | Other modes | 24 | 15.5 |
| Usage of Oxygen during COVID-19 | Yes | 99 | 63.8 |
|  | No | 56 | 36.2 |
| Usage of steroids during COVID-19 | Yes | 142 | 91.6 |
|  | No | 13 | 8.4 |
| DM type 2 | Yes | 122 | 78.7 |
|  | No | 33 | 21.3 |
| Duration from diagnosis of COVID to admission for mucormycosis (days) | Median (IQR) | 22 (15.0 - 31.0) |  |

### Classification of Cases as per Case Definition

The cases in the present study were classified as either proven or possible mucormycosis (Table 3). Those undergoing surgery and had a histopathological and/or culture evidence of mucormycosis, labelled as proven mucormycosis, were 125 (80.6%). Of these, 116 (74.8%) had evidence of mucormycosis with Rhizopus arrhizus being the commonest organism isolated and another 9 cases (5.8%) had evidence of mucormycosis with Aspergillus (aspergillus flavus and Aspergillus niger being equally present). Cases labelled as possible mucormycosis on the basis of history of usage of steroids, presence of diabetes mellitus and clinicoradiological findings of rhinosinus disease, but no histopathological/culture confirmation were 30 (19.4%). These included 10 cases (6.5%) where the biopsy specimen obtained from surgery could not confirm the presence of mucor and another 20 cases (12.9%) where surgery was not performed as per the decision of the treating surgeon.

**Table 3.**
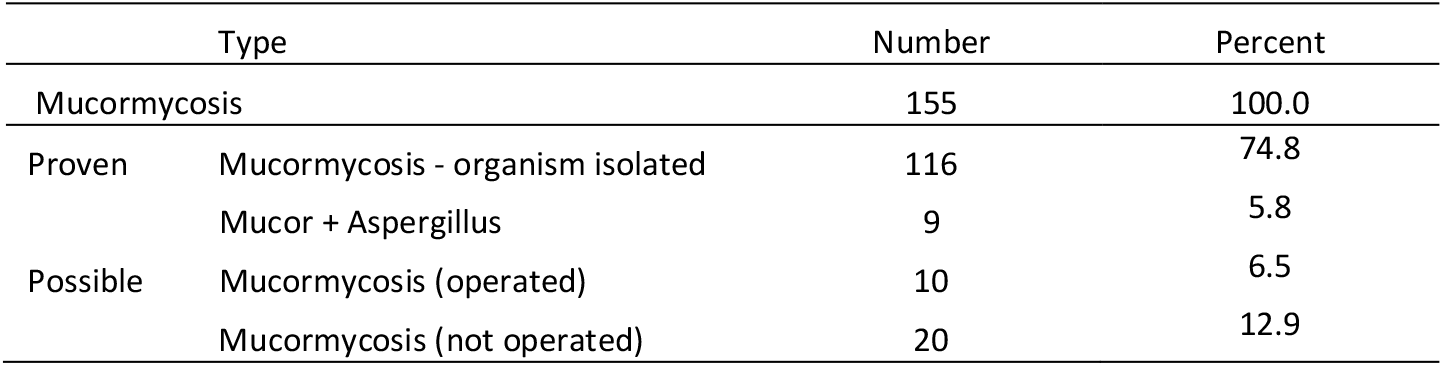
Types of mucormycosis cases in the study.

### Mortality and its Correlates

A total of 26 (16.8%) cases died and others were discharged. The death rate steeply increased from 9.8% in the age-group less than 45 years to 40.0% in the age group 75 years or more (Table 4). The Fisher exact test for overall association between age and death rate was not significant (P = 0.210) but for trend of increasing mortality with age was significant (P = 0.041).

**Table 4.** Differential characteristics of CAM patients in relation to survival and death.

| Characteristics | Total | Survived |  | Died |  | P-Value |
| --- | --- | --- | --- | --- | --- | --- |
|  | n | n | Percentage | n | Percentage |  |
| Total cases | 155 | 129 | 83.2 | 26 | 16.8 |  |
| Age (years) |  |  |  |  |  |  |
| <45 | 41 | 37 | 90.2 | 4 | 9.8 | 0.041 |
| 45-59 | 67 | 56 | 83.6 | 11 | 16.4 |  |
| 60-74 | 42 | 33 | 78.6 | 9 | 21.4 |  |
| ≥75 | 5 | 3 | 60.0 | 2 | 40.0 |  |
| Diabetes Mellitus |  |  |  |  |  |  |
| Yes | 122 | 100 | 82.0 | 22 | 18.0 | 0.42 |
| No | 33 | 29 | 87.9 | 4 | 12.1 |  |
| Steroid usage |  |  |  |  |  |  |
| Yes | 142 | 118 | 83.1 | 24 | 16.9 | 0.889 |
| No | 13 | 11 | 84.6 | 2 | 15.4 |  |
| Oxygen requirement |  |  |  |  |  |  |
| Yes | 45 | 25 | 55.6 | 20 | 44.4 | <0.001 |
| No | 110 | 104 | 94.5 | 6 | 5.5 |  |

| Mode of Oxygen delivery |  |  |  |  |  |  |
| --- | --- | --- | --- | --- | --- | --- |
| NP/FM | 15 | 14 | 93.3 | 1 | 6.7 | <0.001 |
| NRBM | 7 | 5 | 71.4 | 2 | 28.6 |  |
| NIV | 2 | 1 | 50.0 | 1 | 50.0 |  |
| MV | 21 | 5 | 23.8 | 16 | 76.2 |  |
| Place of treatment in hospital |  |  |  |  |  |  |
| Ward | 110 | 100 | 90.9 | 10 | 9.1 | <0.001 |
| ICU | 45 | 29 | 64.4 | 16 | 35.6 |  |
| Laboratory Parameters |  |  |  |  |  |  |
| CRP Units<br>Median (IQR) | 98 | 77 | 40.3<br>(16.3 - 115.3) | 21 | 143.4<br>(56.8 - 216.1) | <0.001 |
| Ferritin Units<br>Median (IQR) | 43 | 30 | 615.65<br>(342.57 - 1273.32) | 13 | 1063.0<br>(693.8 - 1632.0) | 0.048 |
| HbA1c% Median<br>(IQR) | 102 | 83 | 9.8 (8.7 - 11.5) | 19 | 9.8 (7.7 - 10.7) | 0.873 |
| < 8 | 20 | 17 | 85 | 3 | 15 | 0.626 |
| ≥ 8 | 82 | 65 | 79.2 | 16 | 20.7 |  |
| Treatment modalities |  |  |  |  |  |  |
| Surgery+Antifungals | 135 | 119 | 88.1 | 16 | 11.9 | <0.001 |
| Antifungals alone | 20 | 10 | 50.0 | 10 | 50.0 |  |
| Ampho B | 127 | 107 | 84.3 | 20 | 15.7 |  |
| Posaconazole | 105 | 92 | 87.6 | 13 | 12.4 |  |
| Antibiotics | 140 | 118 | 84.3 | 22 | 15.7 |  |

Death rates in various other subgroups are also in Table 4. Mortality was particularly high in the cases requiring oxygen (with highest mortality reaching 76.2% in those on mechanical ventilator). The mortality was only 9.1% in those who remained in the ward and more than three times (35.6%) in those in ICU (P < 0.001).

Median CRP level in those who died was more than three times of the level in those who survived (Table 4). Median ferritin level was more than one-and-a-half times. Both these differences were statistically significant (P < 0.05).

In our study, the presence of diabetes mellitus, though associated with a higher mortality (18.0%) as compared to nondiabetics (12.1%), the difference was not statistically significant (P = 0.420). The median HbA1c, calculated from 102 values that were available, were also not correlated with the mortality (P = 0.873, Table 4). Using HbA1c above 8% as a measure of poorly controlled diabetes, the mortality in those with HbA1c <8% was 15.0% and for those with HbA1c ≥ 8% it was 20.7% but again not statistically significant (P = 0.626).

Combined modality treatment (antifungals plus surgery) significantly reduced mortality (16 deaths out of 135 cases) (11.9%) compared to only medical management with antifungals (10 deaths out of 20 cases) (50.0%) (P<0.001) (Table 4).

The part affected was classified as per the clinical, radiological, and operative findings (Table 5). Most (71, 45.8%) had disease limited to nose and sinuses (Rhino mucormycosis, RM), 47 patients (30.3%) had eye, nose, and sinus involvement (Rhino-ocular mucormycosis, ROM), and 33 patients (21.3%) had brain involvement along with nose, sinus and eye involvement (Rhino-oculo-cerebral mucormycosis, ROCM). Four patients (2.6%) had disseminated mucormycosis as they had involvement of two non-contiguous areas (lungs plus sinuses with or without eye). The mortality in RM was 9.9%, in ROM was 23.4%, in ROCM was 18.2%, and in DM was 50% with a P-value just reaching significance at 0.058, suggesting significantly increased mortality with more aggressive disease.

**Table 5.** Mortality in relation to extent of mucormycosis.

| Extent of Disease | Total Cases |  | Survived |  | Died |  | P-value |
| --- | --- | --- | --- | --- | --- | --- | --- |
|  | n | Percentage | n | Percentage | n | Percentage |  |
| Rhino mucormycosis (RM) | 71 | 45.8 | 64 | 90.1 | 7 | 9.9 | 0.058 |
| Rhino ocular mucormycosis (ROM) | 47 | 30.3 | 36 | 76.6 | 11 | 23.4 |  |
| Rhino ocular cerebral mucormycosis (ROCM) | 33 | 21.3 | 27 | 81.8 | 6 | 18.2 |  |
| Disseminated Mucormycosis (DM) (sinus plus pulmonary) | 4 | 2.6 | 2 | 50.0 | 2 | 50.0 |  |

## Discussion

Mucormycosis is not a new entity but the sudden unpredictable rapid rise in a COVID-19 era made it a different entity and has been known as COVID-19 associated mucormycosis (CAM). Popularly referred to as black fungus, mucormycosis is caused by a group of moulds called mucormycetes, and is a potentially fatal infection if inadequately treated [6]. India contributed to approximately 71% of the global cases of CAM based on the published literature from December 2019 to the start of April 2021 [7].

As many as 91.6% patients in our study reported use of steroids during their COVID-19 treatment and 63% received oxygen at that time. More than three-fourths (78.7%) had diabetes mellitus, type-2. Patel et al. [8] observed the presence of diabetes mellitus in 73.5% of their patients, malignancy in 9.0%, and history of transplant in 7.7% of patients. These three constituted major risk factors for the occurrence of mucormycosis in their series. Ravani et al. [9] reported that the major risk factor for mucormycosis was uncontrolled diabetes Mellitus (96.7%) and COVID-19 illness with the use of steroids (61.2%). A data of 18 patients with aggressive maxillofacial and rhino-cerebral-orbital fungal infections was analysed by Moorthy et al. [10]. All these were COVID-19 positive and16 of them received steroids for COVID-19 management. A review of 41 cases of COVID-19 associated mucormycosis conducted by John et al. found that 94% cases had diabetes mellitus as comorbidity with mean HbA1c of 10%. Out of these, 25 developed mucormycosis following recovery from COVID-19 and all except 1 received systemic steroids for management of COVID-1 while 3 received tocilizumab. Pal et al. [11], in their systemic review, reported overall prevalence of diabetes to be 85% (Diabetes mellitus, type-2 in 79% including new-onset in 16%, Diabetes mellitus type-1 in 6% cases). Nearly 29% in their study had presence of ketoacidosis. Similarly, 85% patients reported usage of glucocorticoids during COVID-19 treatment and 87% reported use of broad-spectrum antibiotics.

During acute COVID-19 disease, more than one-fourth (26.4%) cases in our series were treated at home and the remaining (73.6%) needed hospitalisation. Eighteen cases (11.6%) had concurrent CAM, while the rest (88.4%) got admitted after a median interval of 22 days (IQR 15-31 days), with the longest interval being 69 days from the diagnosis of COVID-19. Mucormycosis occurrence has been described as late as 42 days and 90 days following COVID-19 [12,13]. This shows how the patients can be affected post-COVID. Pal et al. [11] reported 39% of cases with concurrent CAM while the remaining 61% had a mean interval of 22 days between the two illnesses.

We observed in our study that most (71, 45.8%) of the patients had disease limited to nose and sinuses (Rhino mucormycosis, RM), 47 patients (30.3%) had eye, nose and sinus involvement (Rhino-ocular mucormycosis, ROM), and 33 patients (21.3%) had brain involvement along with nose, sinus and eye involvement (Rhino-oculo-cerebral mucormycosis, ROCM). Four patients (2.6%) had disseminated mucormycosis as they had involvement of two non-contiguous areas (lungs plus sinuses with or without eye). A multicentric observational study on 465 patients of mucormycosis from India conducted by Patel et al. reported the presence of rhino orbital mucormycosis in 67.7% cases, pulmonary mucormycosis in 13.3%, and cutaneous mucormycosis in 10.5%. The study by John et al. found that 41% of their patients had rhino-orbital, 27% had rhino-orbital-cerebral, 7% had rhino cerebral, 7% had sinusitis alone and 7% had pulmonary mucormycosis. Pal et al., in their systemic review, reported following site specific mucor: Rhino-orbital (42%), nose/paranasal sinus (16%), rhino-cerebral (4%), rhino-orbito-cerebral (24%), pulmonary (10%), and the rest 1% each (oral, gastrointestinal, cutaneous, and disseminated). Thus, there are varied findings in different studies.

In our study, 74.8% cases had evidence of mucormycosis with Rhizopus arrhizus being the commonest organism isolated and another 5.8% had evidence of mucormycosis with Aspergillus (aspergillus flavus and Aspergillus niger being equally present). In 6.5% cases the biopsy specimen obtained from surgery could not confirm the presence of mucor and in another 12.9% surgery was not performed as per the decision of the treating surgeon. These 19.3% cases were labelled as ‘possible’ mucormycosis. Patel et al. in their study were able to identify the fungus species in the specimen of 290 cases that showed the growth of rhizopus in 231 (79.7%) cases. Moorthy et al., on examination for species of fungus, found 16 out of a total of 18 cases had mucormycosis while 1 had aspergillus and 1 case had mixed growth on histopathology specimen. In the systematic review of 99 cases by Pal et al, data on causative Mucorale was available in 21 patients only: 85% had Rhizopus sp, 10% had Mucor sp and 5% had Lichtheimia sp. Thus, there was no pattern across studies.

In our cohort, surgery was performed in 135 (87.1%) cases, predominantly functional endoscopic sinus surgery (FESS) (83.9%). All these patients received antifungals also. Many had multiple surgeries. Orbital decompression (13.5%), exenteration (4.5%), maxillectomy (7.1%) and craniotomy (1.3%) were the other surgical procedures performed in these patients. In our series, a total of 127 patients (81.9%) received Amphotericin B and 105 patients (67.7%) received Posaconazole (many as combination therapy) and 140 patients (90.3%) received antibiotics. In the study by Patel et al., management of mucormycosis with amphotericin B as primary treatment was done in 81.9% of cases while 11.4% received combination therapy with posaconazole. Surgery was performed only in 62.2% of cases, with the surgical rate being higher in cases of rhino orbital mucormycosis. Pal et al. found the use of adjuvant surgery for management to have been undertaken in 81% of patients. Ravani et al. reported treatment with Liposomal Amphotericin B was done with an average duration of 18.93 days and exenteration was done in 12.9% of cases.

The overall mortality in our study was 16.8%. The death rate steeply increased from 9.8% in the age-group less than 45 years to 40.0% in the age group 75 years or more. Mortality was particularly high in the cases requiring oxygen with highest mortality reaching 76.2% in those on mechanical ventilator. In our study, the presence of diabetes mellitus, though associated with a higher mortality (18.0%) as compared to non-diabetics (12.1%), the difference was not statistically significant (P = 0.420). The mortality in those with HbA1c <8% was 15.0% and for those with HbA1c ≥ 8% it was 20.7% but again not statistically significant (P = 0.626). Combined modality treatment (antifungals plus surgery) significantly reduced mortality (16 deaths out of 135 cases) (11.9%) compared to only medical management with antifungals (10 deaths out of 20 cases) (50.0%) (P<0.001). The mortality in RM was 11%, in ROM was 26%, in ROCM was 18%, and in DM was 50% with a P-value just reaching significance at 0.058, suggesting significantly increased mortality with more aggressive disease. Treatment by different drugs (amphotericin B, posaconazole, and antibiotics) did not make much of a difference in mortality. Patel et al. reported overall better outcomes in cases that received both medical and surgical management. 90-day mortality was reported in 52 % of cases, which is much higher than in our cases (16.8%). The duration of symptoms before hospitalization, the site of involvement, and treatment with deoxycholate Amphotericin B preparation were associated with the increased mortality. They did not report any statistically significant survival difference in diabetics (P=0.67) or those with uncontrolled diabetes (P=0.66) and ketoacidosis (P=0.26) Pal et al. reported adjuvant surgery for management in 81% of patients and found this associated with better clinical outcomes. They also did not find any statistically significant difference in the survival between diabetics, as there were 88% diabetics amongst those who survived and 76% among those who died (P=0.161). Ravani et al. reported cerebral involvement and HbA1c value of ≥8 as the most important predicting factor for survival in cases of mucormycosis. Our findings do not confirm this.

In our earlier study [14] on comparison of the clinical characteristics between the cases in two waves of COVID-19 in the same group of hospitals in North India, the mortality rate was 10.5% during Wave-2 in the hospitalized patients. The mortality reported in patients of CAM, in the present study was 16.8%, which is one-and-a-half times. Clearly, presence of secondary infections, especially mucormycosis, increased the mortality. In the same study, we reported prevalence of diabetes to be 44.9% in the hospitalised COVID-19 patients. In patients who subsequently developed CAM, as per our present study, this was 78.7%. The usage of steroids was 87% and 91.6%, respectively. This comparative between two closely matched patient populations over similar time frame (March to July 2021), clearly identifies diabetes and use of steroids, as important risk factors for development of mucormycosis, And once mucormycosis complicates COVID-19, the mortality goes up significantly.

## Conclusions

COVID-19 associated mucormycosis has become a disease of epidemic proportions in several Indian states during the second wave of COVID-19 pandemic. In the present study, we could identify diabetes and use of steroids, as clear risk factors. Although India always has had highest reported numbers of mucormycosis in the world, physicians never reported mucormycosis as much as occurred during COVID-19 wave-2 in the country. There is a need to explore other causations for CAM, including direct role played by SARS-CoV-2. Timely diagnosis by having a high index of suspicion and combined early surgical and medical management, offers the better chance of survival in an otherwise fatal disease.

## Data Availability

This is a retrospective, observational, multi-centre study that included all the cases recorded as mucormycosis either at discharge or at death, between 1st March 2021 and 15th July 2021 in our network of hospitals in North India. Their records were retrieved from the Electronic Health Records system and their demographic and clinical profile, the hospital course, and the outcome were noted.

## Contributions of authors

SB designed the study concept, finalized the draft, and contributed patients for the study, AI did the statistical analysis and contributed to the draft, MA wrote the initial manuscript, MM contributed in manuscript writing and contributed cases, VJ contributed in concept designing, data collection and manuscript writing, rest all are clinicians (ENT, Internal Medicine, Pulmonology and Critical Care)and contributed and treated patients in the present study.

## ACKNOWLEDGMENTS

We wish to acknowledge and thank Taruna Sharma, Suraj Singh Bisht, Jaya Vohra, Lokesh Kumar, Adil Chaudhary, Meena Jaswal, Pawan Sharma, Shivani Rawat, Biswasi Tirkey, Arnav Jha, Dr. Istiak Mondal and Dr. Aditya Jain, for their help at various stages of the study.

